# Twin pair analysis uncovers novel links between DNA methylation, mitochondrial DNA quantity and obesity

**DOI:** 10.1101/2024.04.02.24304959

**Authors:** Aino Heikkinen, Vivienne F C Esser, Sara Lundgren, Seung Hyuk T Lee, Antti Hakkarainen, Jesper Lundbom, Juho Kuula, Per-Henrik Groop, Sini Heinonen, Päivi Pajukanta, Jaakko Kaprio, Kirsi H Pietiläinen, Shuai Li, Miina Ollikainen

**Author notes:** **Corresponding author:** Aino Heikkinen ( /).

## Abstract

Alterations in mitochondrial metabolism in obesity may indicate disrupted communication between mitochondria and nucleus, crucial for adapting to changing metabolic demands. Epigenetic modifications, particularly DNA methylation, may influence this intricate interplay, though the specifics remain poorly understood. Leveraging data from the subcohort of the Finnish Twin Cohort (n=173; 86 full twin pairs) that includes comprehensive measurements of obesity-related outcomes, mitochondrial DNA quantity (mtDNAq) and nuclear DNA methylation levels in adipose and muscle tissue, we identified one locus at *SH3BP4* (cg19998400) significantly associated with mtDNAq in adipose tissue (FDR<0.05). *SH3BP4* methylation correlated with its gene expression. Additionally, 14 out of the 35 obesity-related traits displayed significant associations with both *SH3BP4* methylation and mtDNAq in adipose tissue. Using the method that infers causality from examination of familial confounding (ICE FALCON) our data suggests that mtDNAq, insulin sensitivity and certain body fat measures are causal to *SH3BP4* methylation. The examination of mtDNAq and obesity-related traits suggested causation from mtDNAq to obesity which could not, however, be distinguished from potential unmeasured within-individual confounding. In conclusion, our findings underscore the impact of mtDNAq on DNA methylation and expression of the *SH3BP4* gene within adipose tissue, with potential implications for obesity.

## Introduction

The significance of mitochondria as a primary energy source for cell growth and survival is indisputable. While mitochondria contain their own circular 16.6 kb genome, most of the genes required for mitochondrial function are encoded in the nuclear DNA. Therefore, cells require synchronized communication between the nucleus and mitochondria, known as mitonuclear communication, to adapt to changing metabolic demands. This intricate interplay can be subject to regulation by epigenetic mechanisms^1^, including DNA methylation^2^, which may influence the activity of gene expression. Consequently, the importance of DNA methylation for mitochondrial function has been highlighted in the literature^3^.

The existing body of literature supports a bidirectional relationship between mitochondrial function and nuclear DNA methylation. On one hand, different mitochondrial haplotypes and variants can lead to differences in DNA methylation^4–7^ via numerous metabolic pathways, such as the methionine cycle and the production of methyl groups^3^. On the other hand, nuclear DNA methylation may impact mitochondrial metabolism by regulating mitochondrial-associated gene expression^8,9^. The term ‘mitochondrial metabolism’ is used here as an umbrella term for any measurable trait associated with mitochondrial metabolism such as mtDNA quantity (mtDNAq), copy number, biogenesis, dynamics, and OXPHOS (oxidative phosphorylation) activity.

The vital role of mitochondria in cellular energy metabolism has placed them in the center of interest in human traits and diseases with metabolic symptoms, including obesity. Obesity is characterized by excess body weight which can lead to a range of systemic metabolic disturbances, and it has been previously associated with compromised mitochondrial biogenesis and OXPHOS capacity^10,11^. Furthermore, obesity is a highly heterogeneous trait, and the precise molecular phenotypes contributing to the altered mitochondrial metabolism remain unclear. Additionally, the exact tissue-specific roles of mitochondria in obesity are not well documented, although it has been shown that adipose tissue metabolism may be more affected by the acquired weight than muscle tissue^11^.

The research on the role of DNA methylation underlying obesity-associated decline in mitochondrial metabolism remains scarce. A recent study showed that mtDNAq influences cardiovascular disease and mortality through changes in DNA methylation^12^. DNA methylation profiles of adipocyte progenitor cells of individuals with obesity have also been linked to mitochondrial metabolism^13^. Despite these insights, the causal pathways and comprehensive characterization of the orchestrated effects of DNA methylation and mitochondrial decline in obesity and obesity-related phenotypes remain elusive.

Here, we aimed to identify DNA methylation sites associated with differences in mtDNAq in two primary tissues affected by obesity, namely subcutaneous adipose tissue and skeletal muscle. Additionally, we explored whether these methylation sites are linked to a comprehensive range of obesity-related outcomes and other related phenotypes, including several anthropometric and body composition measures, clinical biomarkers, physical activity and biological aging^14^. Finally, we explored the potential causal relationships between mtDNAq, DNA methylation and obesity-related measures with a method called Inference about Causation from Examination of FAmilial CONfounding (ICE FALCON)^15^, utilizing the monozygotic (MZ) twin pairs of the study cohort.

## Results

A total of 173 individuals participated in the study, comprising complete 81 MZ twin pairs and 5 DZ twin pairs (Table 1). The age range of the study cohort spanned from 23 to 70 years old, with females accounting for 59% of the cohort (Table 1). The mean BMI and fat percentage were 29.2kg/m^2^ and 39.6%, respectively, indicating that the sample is predominantly with overweight. In addition, average fasting glucose levels (5.6 mmol/l) and

**Table 1.** Participant characteristics.

|  |  |
| --- | --- |
| N (twin pairs, singletons) | 173 (86, 1) |
| MZ / DZ (full twin pairs) | 163 (81) / 10 (5) |
| Age, mean (range) | 45.7 (22.8–69.3) |
| Female, % | 59.0 |
| Smoking, % | Never 39.3<br>Current 30.1<br>Former 30.6 |
| **Obesity-related measures, Mean (range) SD** |  |
| Weight (kg) | 83.7 (48.7–143.6) 18.3 |
| BMI (kg/m<sup>2</sup>) | 29.2 (19.7–45.9) 5.8 |
| Waist circumference (cm) | 97.6 (65.2–144.3) 15.7 |
| WHR | 0.9 (0.8–1.1) 0.1 |
| Fat (%) | 36.6 (8.9–57.3) 9.6 |
| Fat (kg) | 31.9 (7.4–66.0) 13.0 |
| Fat-free mass (kg) | 49.8 (29.5–76.7) 10.8 |
| Intra-abdominal fat (cm<sup>3</sup>) | 1,015 (152–3,950) 870 |
| Subcutaneous fat (cm<sup>3</sup>) | 4,582 (827–15,129) 2,830 |
| Adipocyte volume (mm<sup>3</sup>) | 539 (123–1,131) 218 |
| Liver fat (%) | 2.7 (0.1–22.4) 4.0 |
| Total cholesterol (mmol/l) | 4.8 (2.7–7.7) 0.9 |
| HDL (mmol/l) | 1.5 (0.5–3) 0.5 |
| LDL (mmol/l) | 3 (1–5.1) 0.8 |
| Triglycerides (mmol/l) | 1 (0.3–5.9) 0.6 |
| hsCRP (mg/l) | 2.3 (0.1–9.7) 2.3 |
| Adipsin (µg/ml) | 1.2 (0.7–1.7) 0.2 |
| Adiponectin (ng/ml) | 3,320 (1,310–7,710) 1,408 |
| ALAT (U/l) | 29.3 (14–127) 20.1 |
| ASAT (U/l) | 29.2 (7–131) 14.3 |
| Systolic blood pressure (mmHg) | 133 (97–195) 19 |
| Diastolic blood pressure (mmHg) | 80 (48–110) 78 |
| Fasting glucose (mmol/l) | 5.6 (4.0–16.1) 1.1 |
| Fasting insulin (mU/l) | 8.1 (0.9–34.3) 5.9 |
| HOMA-IR | 2.1 (0.2–9.9) 1.8 |
| Matsuda index | 6.7 (0.7–34.2) 4.9 |
| **Physical activity measures (Baecke scale), Mean (SD)** |  |
| Leisure-time physical activity | 2.9 (0.6) |
| Sports activity | 2.7 (0.9) |
| Work activity | 2.5 (1.0) |
| Total activity | 8.1 (1.5) |

ALAT = Alanine aminotransferase; ASAT = Aspartate aminotransferase; BMI = body mass index; DZ = dizygotic; HOMA-IR = Homeostatic model assessment for insulin resistance; hsCRP = high-sensitivity C-reactive protein; MZ = monozygotic; WHR = waist-to-hip ratio

HOMA-IR measures (2.1 units) were slightly elevated which suggests a reduced insulin sensitivity. Twenty-two participants were diagnosed with type 2 diabetes. The main analysis strategy is described in Figure 1, while Supplementary Table 1 summarizes the epigenetic age acceleration estimates of the study participants. Furthermore, the BMI-discordant MZ twin pairs with adipose tissue data (n=71 pairs) used in the causal inference analysis are presented in Supplementary Table 2.

**Figure 1.**
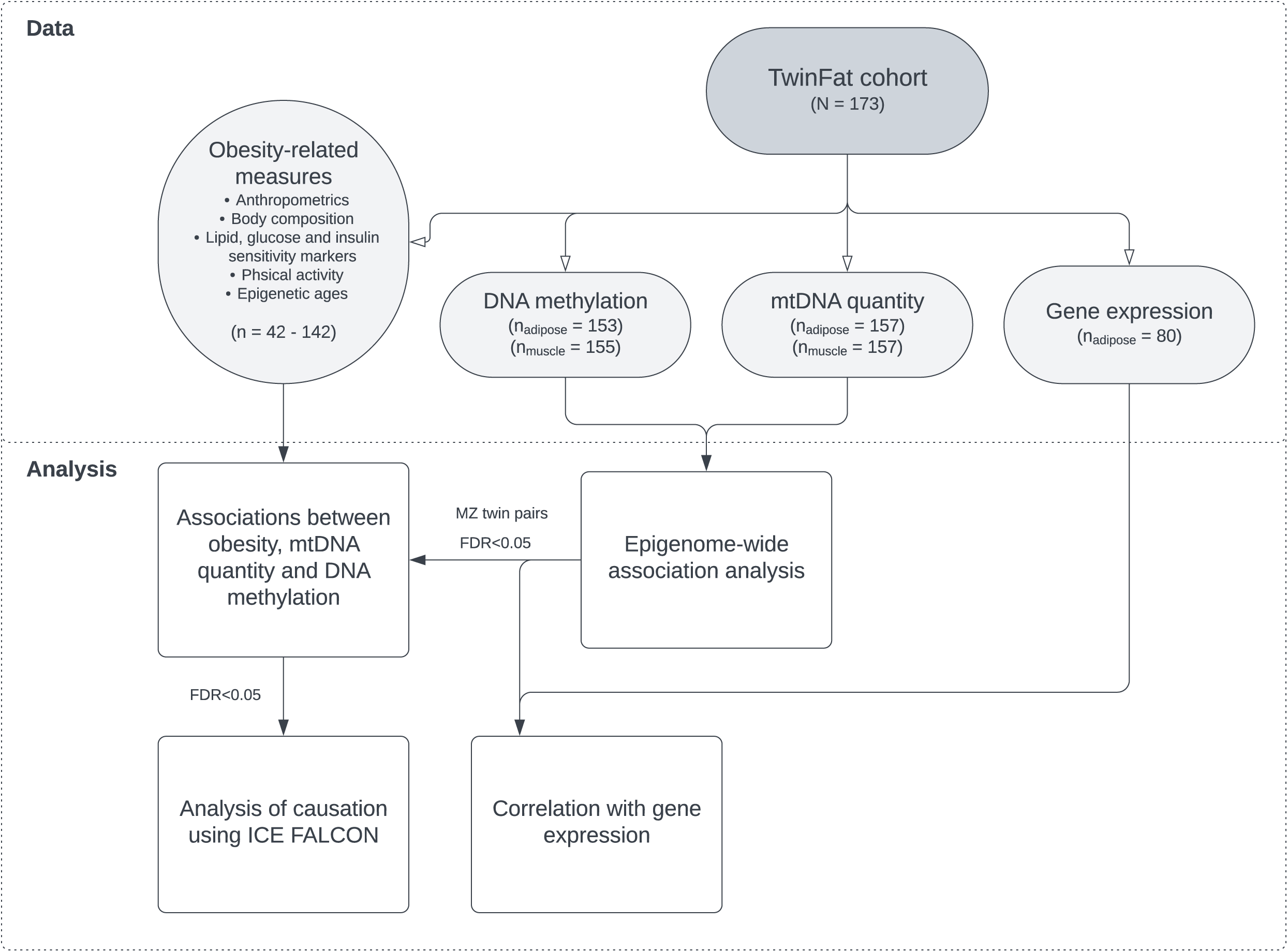
Study flowchart.

### Differential methylation analysis

To investigate whether nuclear DNA methylation was associated with mtDNAq, we performed an epigenome-wide association analysis (EWAS). In adipose tissue (n=153 individuals), we identified one CpG site (*cg19998400*) that was inversely associated with mtDNAq (FDR=0.002) (Fig. 2a). The CpG is in the enhancer region of *SH3BP4* (SH3 domain binding protein 4) gene that codes for a protein involved in intracellular signaling pathways. Another CpG (*cg17468563*) located in the *DHRS3* (Dehydrogenase/Reductase 3) enhancer region showed marginal association with mtDNAq (FDR=0.078) (Fig. 2a). Muscle tissue CpG methylation (n=155 individuals) was not associated with mtDNAq (Fig. 2b). The association profiles of the two tissues did not show similar patterns, as none of the top 100 CpG were found to be in common between adipose and muscle samples. Additionally, there was no strong correlation between the regression effect sizes (Supplementary Fig. 1). The rest of the manuscript focuses explicitly on the findings in adipose tissue, given that we did not detect any statistically significant mtDNAq-linked DNA methylation sites in muscle.

**Figure 2.**
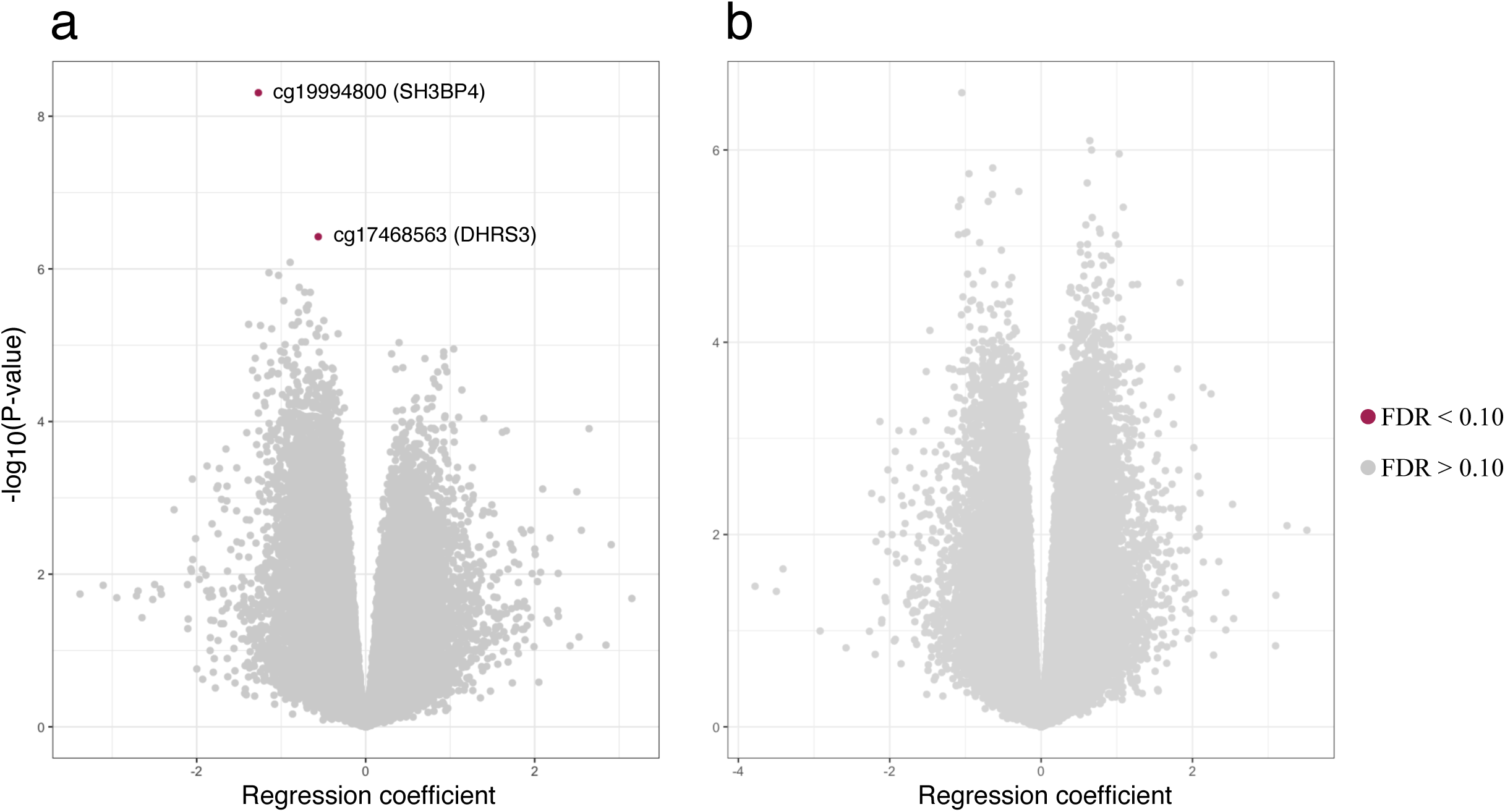
Volcano plot of the epigenome-wide association study on mtDNA quantity in (a) adipose tissue (n=153 individuals) and (b) skeletal muscle (n=155 individuals). Red dots indicate CpGs with FDR<0.10.

### Causal inference between adipose tissue mitochondrial DNA quantity and identified CpG sites

To investigate the potential evidence for causation underlying the associations between mtDNAq and the two identified CpG sites in adipose tissue, we performed ICE FALCON analysis for the complete MZ twin pairs with data on adipose tissue DNA methylation (n=68 MZ pairs). We analyzed each CpG separately given that we cannot assume them to have identical causal pathways. For instance, the *cg17468563* has been reported to be under genetic control of multiple loci^16^ whereas no meQTLs have been identified for *cg19998400*. Our data was consistent with the hypothesis that mtDNAq is causally linked to cg*19998400* (Table 2). This was suggested when we set mtDNAq as the independent variable and observed a marginal cross-twin cross-trait association (Model 2: B_cotwin_=-0.160, p =0.091) which was attenuated towards null when adjusting for the twin’s own mtDNAq. There was no evidence of change in the within-individual association. Conversely, when using mtDNAq as the dependent variable, we did not observe any cross-twin cross-trait associations in Models 2 or 3. However, the increased absolute value of the regression coefficient in Model 3 is in line with mtDNAq being causal to methylation at *cg19998400* (Table 2). The change was not statistically significant, which may reflect a low sample size and consequently reduced statistical power. The results from ICE FALCON on *cg17468563* were more ambiguous, with the regression coefficients pointing to either causation from mtDNAq to methylation at *cg17468563* or presence of within-individual confounding (Table 2).

**Table 2.** Results from ICE FALCON analysis between mtDNA quantity and the identified CpG sites (n = 68 monozygotic twin pairs) in adipose tissue. Regression models were adjusted for age, sex, smoking, BMI and methylation beadchip and row. P-values < 0.05 are bolded.

| Formula<br>(Y/X<br>variable) | Coef*<br>($\beta_{self}$ /<br>$\beta_{cotwin}$) | Model 1 | | | Model 2 | | | Model 3 | | | Change in<br>coefficients | |
| --- | --- | --- | --- | --- | --- | --- | --- | --- | --- | --- | --- | --- |
|  |  | Est | SE | P | Est | SE | P | Est | SE | P | Est | P |
| mtDNAq (Y)<br>cg19998400 (X) | $\beta_{self}$ | -0.350 | 0.061 | **9.2E-09** | | | | -0.358 | 0.060 | **2.1E-09** | -0.008 | 0.728 |
| | $\beta_{cotwin}$ | | | | 0.006 | 0.070 | 0.930 | -0.067 | 0.059 | 0.257 | -0.073 | 0.426 |
| cg19998400 (Y)<br>mtDNAq (X) | $\beta_{self}$ | -0.487 | 0.098 | **7.3E-07** | | | | -0.475 | 0.102 | **3.6E-06** | 0.013 | 0.643 |
| | $\beta_{cotwin}$ | | | | -0.167 | 0.081 | **0.039** | -0.055 | 0.072 | 0.444 | 0.112 | 0.351 |
| mtDNAq (Y)<br>cg17468563 (X) | $\beta_{self}$ | -0.431 | 0.092 | **2.9E-06** | | | | -0.428 | 0.092 | **3.6E-06** | 0.003 | 0.942 |
| | $\beta_{cotwin}$ | | | | 0.163 | 0.080 | **0.043** | 0.143 | 0.073 | 0.051 | -0.020 | 0.788 |
| cg17468563 (Y)<br>mtDNAq (X) | $\beta_{self}$ | -0.342 | 0.082 | **2.9E-05** | | | | -0.331 | 0.082 | **5.0E-05** | 0.011 | 0.664 |
| | $\beta_{cotwin}$ | | | | 0.132 | 0.078 | 0.090 | 0.050 | 0.067 | 0.460 | -0.082 | 0.289 |

\*Standardized regression coefficient; $\beta_{self}$ represents the association between twin's own X and Y variables whereas $\beta_{cotwin}$ is the cross-twin cross-trait association i.e. the association between twin's own X variable with their co-twin's Y variable.
mtDNAq = mitochondrial DNA quantity

### Causal inference between adipose tissue DNA methylation and gene expression

As DNA methylation is a potential mechanism to regulate gene expression, we investigated whether the DNA methylation at *cg19998400* and *cg17468563* were associated with the expression of their respective genes, *SH3BP4* (n=80 individuals) and *DHRS3* (n=80 individuals) in adipose tissue. There was a positive correlation between the expression and methylation of *SH3BP* (r_Pearson_=0.46, p<0.001) (Fig. 3a), where the methylation explained solely 18% of the variation (marginal R-squared) and with familial confounding 58% (conditional R-squared) of the variation in gene expression (Fig. 3c). For the *DHRS3* expression and methylation, we observed a negative correlation (r_Pearson_=-0.66, p<0.001) (Fig. 3b), with a marginal R-squared of 42% and conditional R-squared 56% (Fig 3c).

**Figure 3.**
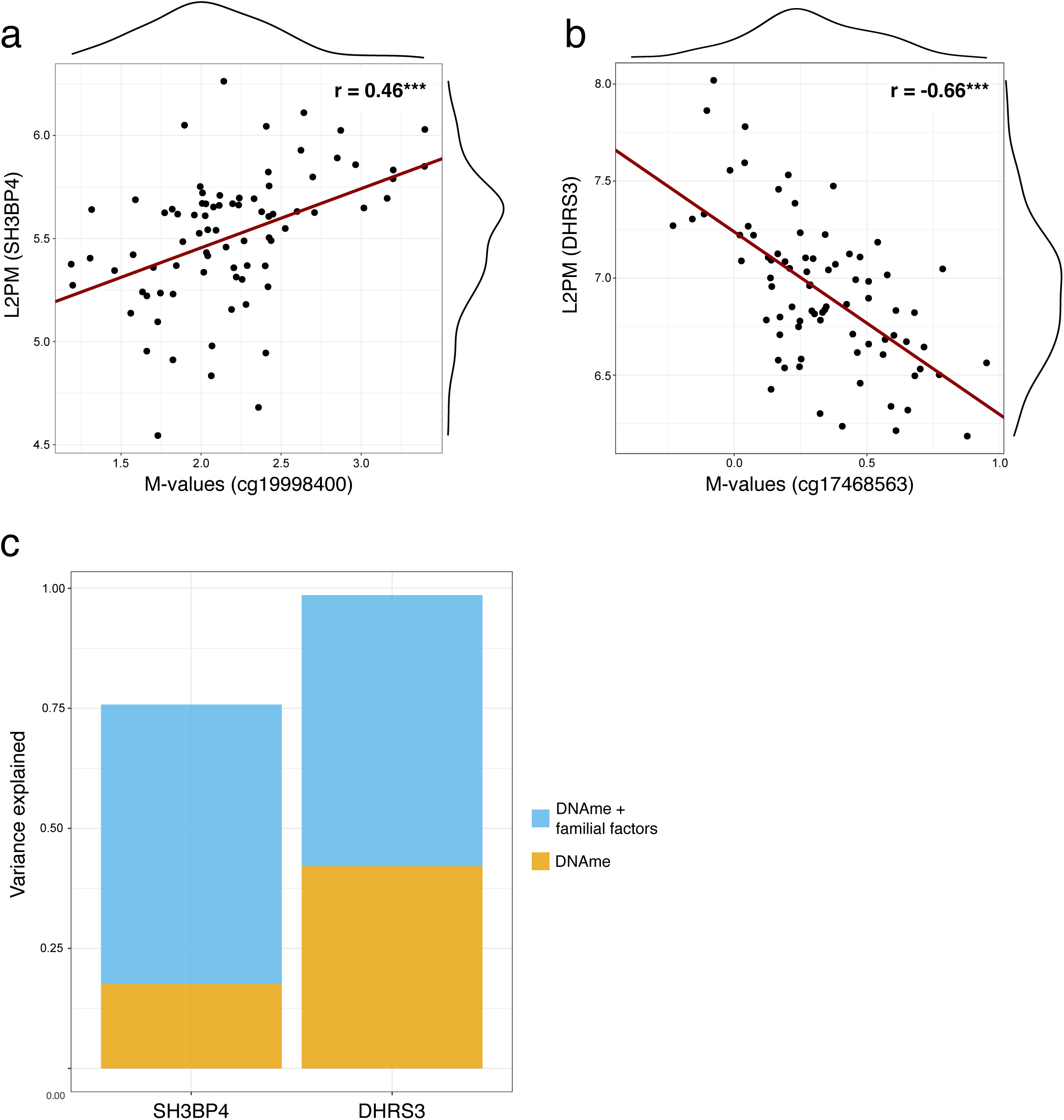
Relationship between DNA methylation and gene expression at two genomic loci in adipose tissue. (a) Correlation between methylation at *cg19998400* and *SH3BP4* expression (n=80 individuals). (b) Correlation between methylation at *cg17468563* and *DHRS3* expression (n=80 individuals). (c) Variation in *SH3BP4* and *DHRS3* expression explained by methylation at *cg19998400* and *cg17468563* (yellow bars), respectively, and combined with common familial factors (genetic or environmental) (blue bars) (n=80 individuals). DNAme = DNA methylation; L2PM = log_2_-counts per million; r = Pearson correlation coefficient

It is important to acknowledge that changes in gene expression patterns can also directly influence DNA methylation levels. Considering this, we conducted an ICE FALCON analysis to explore the direction of causality between gene expression and DNA methylation. We observed a marginal cross-twin cross-trait association between *SH3BP4* expression and *cg19998400* methylation (Model 2: B_cotwin_=-0.112, p =0.078) (Supplementary Table 3) that attenuated towards null when adjusting for twin’s own *cg19998400* levels, whereas the twin’s own regression coefficient remained approximately the same. This observation is consistent with *cg19998400* methylation influencing the *SH3BP4* expression. On the other hand, the possibility for the presence of within-individual confounding cannot be ruled out. In contrast, with *DHRS3* and *cg17468563*, we did not find any compelling evidence for either causation or familial confounding (Supplementary Table 3).

### Obesity-related variables associated with adipose tissue *SH3BP4* methylation and mitochondrial DNA quantity

Considering the well-established connection between mitochondrial dysfunction and obesity, we were interested to study whether the mtDNAq-associated CpG sites would be also associated with different obesity-related variables. We specifically focused on the DNA methylation of *cg19998400* at the *SH3BP4* locus (herein referred to as *SH3BP4* methylation), as our data suggested a potential causal relationship with mtDNAq, in contrast to *cg17468563* methylation at *DHRS3.* Among a set of 35 obesity-related traits, six showed significant association only with mtDNAq, four with *SH3BP4* methylation, and 14 with both mtDNAq and *SH3BP4* methylation (FDR<0.05) (Fig 4., Supplementary Tables 4-5).

**Figure 4.**
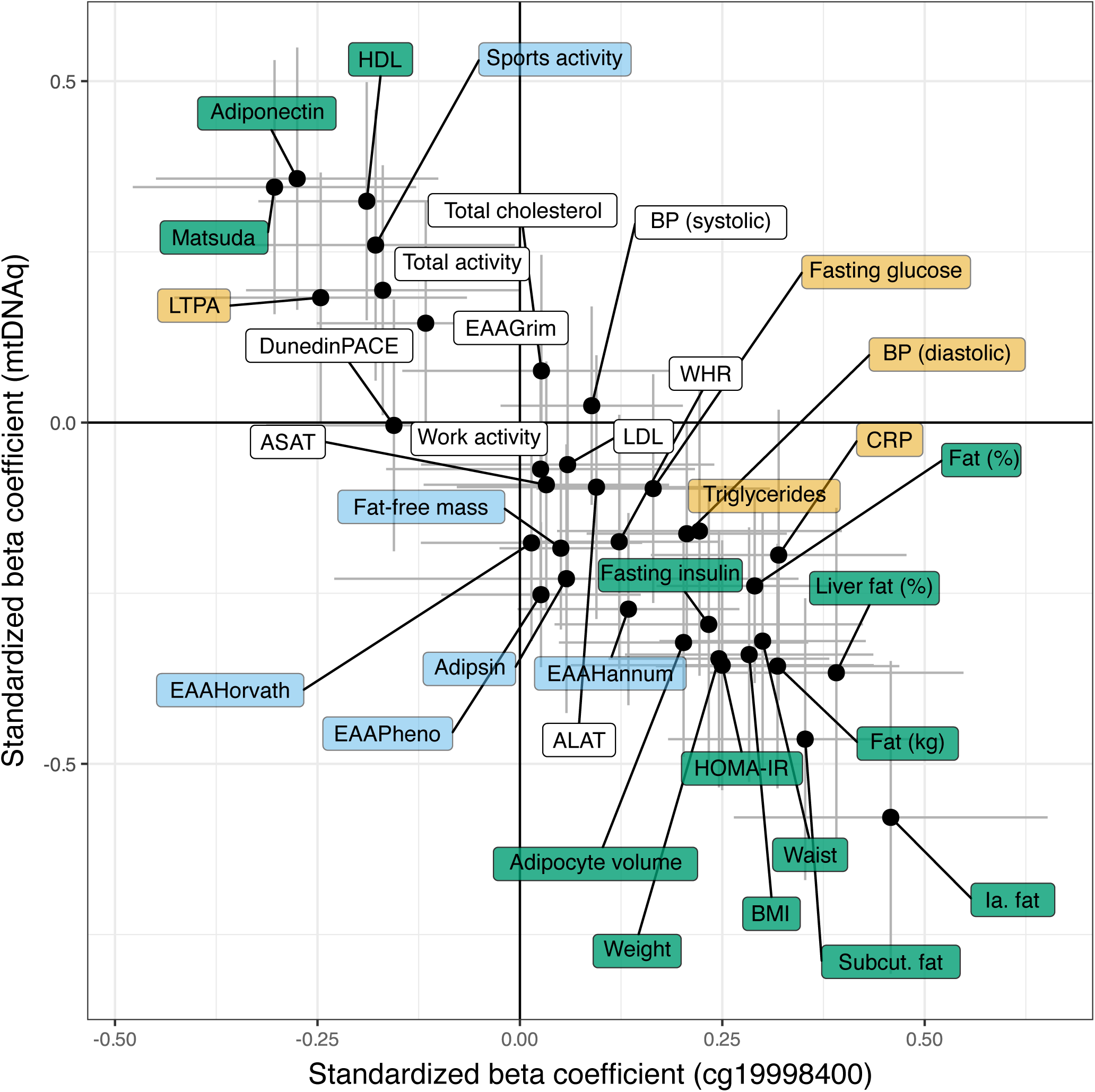
Standardized beta coefficients and standard errors for the associations between obesity-related outcomes and methylation at *cg19998400* (X-axis) or mtDNA quantity (Y-axis) (n=42-142 individuals) in adipose tissue. Yellow background indicates variables with FDR<0.05 associated with *cg19998400* methylation only, blue indicates variables associated with mtDNA quantity only, and green indicates variables associated with both.

Three distinct measures of epigenetic age acceleration (EAA), namely Horvath, Hannum and PhenoAge, displayed a negative association with mtDNAq but not with *SH3BP4* methylation. In addition to EAA, mtDNAq showed negative associations with fat-free mass and adipsin levels, and positive with sports activity. Obesity-related variables that associated exclusively with *SH3BP4* methylation included elevated blood triglyceride levels, hsCRP, fasting glucose levels and systolic blood pressure. The 14 shared associations between mtDNAq and *SH3BP4* methylation included parameters mainly related to body fat composition, insulin sensitivity and HDL cholesterol levels. Consistent with existing literature, higher mtDNAq were correlated with lower body fat, and higher insulin sensitivity, HDL cholesterol levels and adiponectin levels.

### Causal inference between obesity-related traits, and adipose tissue *SH3BP4* methylation and mitochondrial DNA quantity

To discern the potential causal relationship between the 14 obesity-related variables, and both *SH3BP4* methylation and mtDNAq, we employed an ICE FALCON analysis for the complete MZ twin pairs in the cohort (Supplementary Table 2). Our findings indicate that certain variables related to insulin resistance and ectopic fat may exert causal influence on *SH3BP4* methylation, as suggested by significant cross-twin cross-trait association in Model 2 (body fat percentage B_cotwin_=0.170, p=0.074; intra-abdominal fat B_cotwin_=0.235, p =0.031; HOMA-IR B_cotwin_=0.278, p =0.002; Matsuda B_cotwin_=-0.253, p=0.006; fasting insulin B_cotwin_=0.235, p =0.004) that attenuated towards null after conditioning on twin’s own corresponding measures (Fig. 5b). The changes in regression coefficients were significant in fat percentage (p=0.047) and marginally significant in Matsuda (p=0.062). Reversing the regression and using *SH3BP4* methylation as predictor variable also suggested causality from these obesity-related traits to methylation, based on the behavior of the regression coefficients (i.e., the cross-twin cross-trait association increased substantially while the B_self_ remained relatively stable) (Fig. 5c). Results of other obesity-related traits, specifically those measuring body size, liver fat percentage, subcutaneous fat and HDL cholesterol, were less clear. While the behavior of co-twin’s regression coefficients in ICE FALCON may indicate that these variables are a consequence of *SH3BP4* methylation (Fig 5b-c), the opposing signs between Model 1 and Model 2 coefficients, when using *SH3BP4* methylation as predictor variable X, can also suggest the presence of within-individual confounding (Fig 5c). Similarly, the ICE FALCON analysis on the association between mtDNAq and obesity-related variables suggested either mtDNAq being causal to most of the obesity-related variables or being subject to unmeasured within-individual confounding (Fig 5d-e).

**Figure 5.**
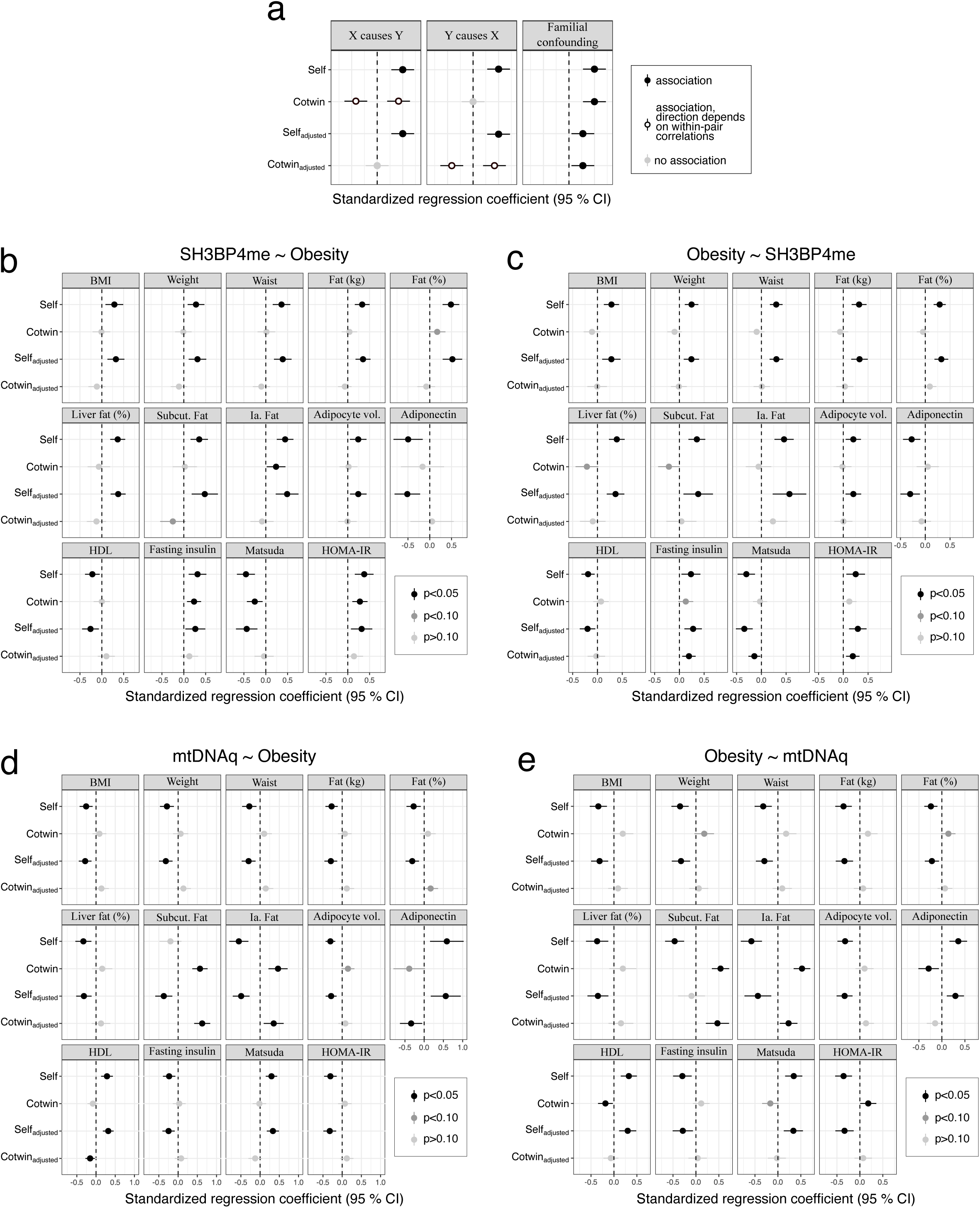
Results from the ICE FALCON analysis between adipose tissue mtDNA quantity, *SH3BP4* methylation and obesity-related outcomes. (a) Behavior of the ICE FALCON regression coefficients of Models 1-3 in each causal scenario. (b-e) Standardized regression coefficients and 95% confidence intervals analysis for the ICE FALCON regression coefficients (n=21-71 pairs) of (b) obesity-related outcomes regressed against *SH3BP4* methylation, (c) *SH3BP4* methylation regressed against obesity-related outcomes, (d) obesity-related outcomes regressed against mtDNA quantity and (e) mtDNA quantity regressed against obesity-related outcomes. ‘Self’ represents the association between twin’s own X and Y variables whereas ‘Cotwin’ is the cross-twin cross-trait association i.e. the association between twin’s own X variable with their co-twin’s Y variable. ‘Adjusted’ refers to the regression coefficients derived from the ICE FALCON Model 3 that includes both twin’s own and their cotwin X variables. HDL = high-density lipoprotein; Subcut. = subcutaneous; Ia. = intra-abdominal

## Discussion

In this study, we identified a significant association between mtDNAq and DNA methylation at *SH3BP4*, which correlated with its gene expression levels. This methylation site was also linked to numerous obesity-related traits, specifically those that measure body fat composition and insulin sensitivity. Leveraging our data on monozygotic twin pairs, we identified potentially causal associations from mtDNAq and obesity-related outcomes to *SH3BP4* methylation in adipose tissue.

The dynamic interplay between mitochondria and the nucleus plays a pivotal role in responding to diverse extracellular signals and metabolic conditions, as suggested by our findings: modifications in mtDNAq may precede changes in *SH3BP4* methylation, which would indicate a retrograde signaling from mitochondria to nuclear DNA methylation. This observation aligns with prior research, although limited, showing the impact of reduced mtDNAq on nuclear DNA methylation in human embryonic kidney cell lines^12^. One of the main hypotheses for retrograde signaling includes the importance of mitochondria in the methionine cycle, and in the production of S-adenosylmethionine. S-adenosylmethionine is a primary methyl donor in cells, which interacts with DNA methyltransferases (DNMT) and therefore can affect DNA methylation^17^. However, the targeted nature of the mitochondria-dependent methylation at specific genomic loci, such as *SH3BP4*, requires further investigation, along with its potential functional implications.

Our study indicated that the DNA methylation of *SH3BP4* and *DHRS3* associated with mtDNAq may be reflected at the gene expression level in adipose tissue. Specifically, methylation of *cg19998400* located at the 5’UTR region of *SH3BP4* correlated positively with *SH3BP4* expression whereas methylation at the gene body (*cg17468563*) of *DHRS3* displayed a negative correlation, which is contrary to the previously observed general pattern of positive association between gene body methylation and expression^18,19^. However, recent research indicates a far more complex relationship between these two factors, heavily influenced by the underlying genomic context^20–22^. Although our study does not pinpoint the molecular mechanisms underlying the association, we underscore the potential implication of these genes and their methylation status in relation to varying mtDNAq in obesity. Moreover, we demonstrated that DNA methylation of both *SH3BP4* and *DHRS3* accounted for a substantial proportion of the variation in the expression of these genes, alongside shared familial factors within the twin pairs. These factors encompass both genetic elements, such as eQTLs, and environmental factors, like age and lifestyle, which cannot be distinguished apart using MZ twin pairs only. Consequently, future studies with the inclusion of a cohort of dizygotic twins or other family members could provide insights into the relative significance of genes and environment.

The link we identified between the mtDNAq-associated methylation site in *SH3BP4* and various obesity-related outcomes, particularly those assessing insulin sensitivity and body fat composition, implies a potential role of mtDNAq-induced methylation in the etiology of obesity. *SH3BP4* acts as a negative regulator in many signaling pathways such as mTORC1 (mammalian target of rapamycin complex 1), a key promoter of cell growth and proliferation^23^, and has been observed to promote adipogenesis, possibly by regulating mitochondrial functions^24–26^. Previous research has demonstrated that mTOR signaling is compromised in obesity, potentially influenced by factors such as diet quality^27–29^ and oxidative stress^30^. Still, the precise contribution of *SH3BP4* methylation and gene expression to cellular function and disease development remains unclear.

Our analysis indicates that changes in DNA methylation at *SH3BP4* may result from alterations in body insulin sensitivity and intra-abdominal fat accumulation. Previous studies conducted on blood have also suggested that methylation may be a consequence rather than a cause of obesity^31,32^. Intriguingly, our findings suggest that only certain obesity-related outcomes, perhaps those more closely tied to metabolic disruptions such as insulin resistance and ectopic fat, are causally associated to *SH3BP4* methylation, while others closely related, such as BMI, are not. This may reflect the high metabolic heterogeneity often observed among people with similar BMI^33^. Nevertheless, the discrepancy warrants further investigation into the pathways that result in changes in *SH3BP4* methylation in the context of obesity.

The connection between mtDNAq and obesity has been established^10,34,35^, yet the exploration of causal pathways has been limited. We identified a dualistic relationship between mtDNAq and obesity-related outcomes which pointed to either causation from mtDNAq to several obesity-related outcomes or the presence of unmeasured within-individual confounding. Excessive nutrient intake is a plausible factor influencing this association, as it is known to impair mitochondrial function^36–38^ and contribute to the excess body weight. Whether nutrient intake serves as a confounder in the association or initiates a pathway mediated by mitochondria leading to obesity remains uncertain. It is also possible that there is a circular relationship between mtDNAq and obesity. Additionally, changes in mtDNAq may manifest only after changes in other mitochondrial parameters, which were not covered in this study, thereby limiting the identification of causal associations between mitochondrial function and obesity, and DNA methylation. Despite this, we demonstrate that both mtDNAq as well as specific obesity-related outcomes may be causal to DNA methylation at *SH3BP4*, via shared or independent molecular pathways.

Aging is widely linked with a decline in mitochondrial metabolism, including reduced mtDNAq^39,40^. We revealed an association between mtDNAq and EAA, measured with Hannum, Horvath and PhenoAge, in adipose tissue, which can indicate that mitochondrial metabolism is one of the key components in driving biological aging. These clocks have been reported to exhibit similar transcriptional signals with one another^41^. While many of the epigenetic clocks were originally developed for whole blood (except for Horvath that is a multi-tissue clock), it has been shown that some of the clocks, including PhenoAge and Hannum, work fairly robustly in other tissues too^41^. Moreover, PhenoAge shows increased age acceleration in cells with depleted mitochondria^41^.

Our study underscores the importance of investigating diseases-affected tissues beyond readily available blood samples. Exploring two primary tissues affected by excess weight, we discovered that the associations between DNA methylation and mtDNAq in obesity are not uniform. Only adipose tissue DNA methylation, but not muscle, was found to be associated with mtDNAq, which may reflect the different roles of these two tissues in obesity. For instance, alterations in adipose tissue seem to be more profoundly associated with metabolic health than those in muscle in obesity^11^. It may be that changes in muscle emerge only after systemic changes, such as insulin resistance, that are followed by the lipid accumulation. It is important to note that our cohort comprised mostly healthy participants, and therefore our findings cannot be necessarily generalized to more severe health complications such as metabolic syndrome or type 2 diabetes. Furthermore, mtDNAq as a proxy for mitochondrial biogenesis may not be directly comparable between the two tissues.

This study encompasses several strengths. First, the carefully phenotyped twin cohort for a comprehensive range of obesity-related outcomes enables disentangling the most significant molecular phenotypes of obesity to mtDNAq. Second, the inclusion of adipose and skeletal muscle tissues broadens the examination of the impact of excess weight and other obesity-related outcomes across tissues. In addition, using monozygotic twins, we can use statistical methods such as ICE FALCON to investigate the causality of the observed associations. The ICE FALCON offers a robust approach to explore the causal relationship in the observational data using related individuals, including twins^15,42–44^ without the need for genetic instrumental variables. To our knowledge, there is a lack of established genetic variants or polygenic scores to estimate mtDNAq specifically in adipose and muscle tissues. This absence of genetic information on mtDNAq prevented us from using statistical methods such as MR-CoD^45^, which depend on genetic data.

However, there are limitations to consider that include the cross-sectional nature of the study, as well as the modest sample size, which substantially limits statistical power. Nevertheless, the uniqueness of our dataset, to our knowledge, sets it apart as the first to integrate DNA methylation and mitochondrial quantity in individuals with obesity using adipose and muscle tissues.

Overall, we demonstrate a potential causal link from adipose tissue mitochondrial metabolism to DNA methylation and expression of *SH3BP4*. Additionally, this connection holds significance in obesity, where certain outcomes related to insulin sensitivity and intra-abdominal fat were seen to contribute to *SH3BP4* methylation, influenced either by mtDNAq or through alternative pathways. We propose the existence of a complex network interconnecting DNA methylation and mitochondrial metabolism in obesity, contributing to the multifaceted nature of obesity as a phenotype. Comprehensive examination of their interplay with various metabolic parameters holds promise for advancing our understanding of the intricate metabolic landscape in obesity.

## Methods

### Study cohort

The study participants originate from the metabolic substudy from the larger Finnish Twin Cohort (FinnTwin12^46^, FinnTwin16^47^ and Older Finnish Twin Cohort^48^). The present substudy was designed to study obesity-related metabolism, and the participants were initially invited to participate based on their self-reported weight and height. Our data consisted of 173 individuals, comprising those with both subcutaneous adipose and muscle tissue data available (n=141), individuals with only subcutaneous adipose tissue (n=16), and those with only skeletal muscle data (n=16) (Table 1).

### Clinical data

The selected study participants were deeply phenotyped for obesity-related clinical measures as described more in detail in van der Kolk et al. 2021^11^. Briefly, anthropometric and body composition were measured after overnight fasting. *Body mass index* (BMI) was calculated from *weight* and *height* (kg/m^2^), measured during on-site visits. In addition, *waist circumference* was measured and *waist-to-hip ratio* (WHR) calculated. *Fat mass*, *fat percentage* and *lean mass* were quantified using dual-energy X-ray absorptiometry. *Intra-abdominal* and *subcutaneous fat volumes* were measured using magnetic resonance imaging (MRI), and *liver fat content* using magnetic resonance spectroscopy (MRS). Supine *blood pressure* measurements were also taken (mean of three measurements).

Blood samples were obtained after overnight fasting, and concentrations of *plasma glucose*, *serum insulin*, *plasma total cholesterol*, *low-density lipoprotein* (LDL), *high-density lipoprotein* (HDL), *triglycerides*, *high sensitivity C-reactive protein* (hsCRP), *alanine aminotransferase* (ALAT) and *aspartate aminotransferase* (ASAT) were measured using standard HUSLAB clinical laboratory procedures. *Homeostatic model assessment-insulin resistance index* (HOMA-IR) and the *Matsuda index* for insulin sensitivity were calculated from the standard 4-point oral glucose tolerance test (OGTT).

Levels of different forms of physical activity (*sport, work, leisure and total*) were assessed using the Baecke questionnaire^49,50^.

### Sample Collection

The samples of this study have been used in previous research^11,51,52^. Briefly, the adipose and muscle tissue samples were collected from subcutaneous abdominal adipose tissue and vastus lateralis muscle, respectively, under local anesthesia (lidocaine). Adipose tissue samples were taken using a surgical technique or needle biopsy, and muscle samples through Bergström needle biopsy. Determination of *adipocyte volume (dm^3^)* has been previously described in Lapatto et al. 2023^52^. The tissue samples for DNA/RNA extraction were snap-frozen in liquid nitrogen.

### Mitochondrial DNA quantity

DNA was extracted from adipose and muscle tissue biopsies using AllPrep DNA/RNA/miRNA Universal Kit (Qiagen). The amount of mtDNA was quantified using quantitative PCR (qPCR), targeting for two mitochondrial encoded genes *ND5* (NADH dehydrogenase 5) and *CYTB* (cytochrome b), and normalized to genomic DNA as measured from *APP* (amyloid-beta precursor protein) and *B2M* (beta-2-microglobulin). Data was processed using the 2^-ΔΔCt^ method with qbase+ software (Biogazelle) to obtain calibrated normalized relative quantities (CNRQ) for the mtDNA quantity. In cases of missing data (due to poor sample quality) for either *ND5* or *CYTB*, we applied a stochastic regression method to input missing values, using the other gene as a reference. Subsequently, we calculated the mean of *ND5* and *CYTB*, which served as the metric for mtDNAq.

### DNA methylation data

High molecular weight DNA was extracted from adipose and muscle biopsies with AllPrep DNA/RNA/miRNA Universal Kit (Qiagen) or QIAmp DNA Mini Kit (Qiagen) and bisulfite converted with an EZ DNA Methylation Kit (ZYMO Research) according to the manufacturers’ protocol. DNA methylation was quantified using Infinium HumanMethylation450K (adipose tissue) or HumanMethylationEPIC BeadChip arrays (adipose and muscle tissues).

DNA methylation data was preprocessed and normalized with R package *meffil*^53^. Due to the modest sample size, adipose tissue 450K and EPIC data were preprocessed together, omitting the platform-specific probes from the analysis. After background and bias correction, we excluded bad quality samples with following criteria: i) Median difference in X and Y chromosome intensities > 3 standard deviations (SDs), ii) Median methylated vs. unmethylated intensity > 3 SDs, iii) unreliable control probes, iv) detection p-value >0.01 in more than 20% of probes and v) bead number < 3 in more than 20% of the probes.

We then applied quantile normalization to reduce technical variation by adjusting for methylation sample slide and control probe PCs. Number of PCs included was estimated from a scree plot separately for adipose and muscle data (Supplementary Fig. 2). Bad quality probes were removed with following criteria: i) Detection p-value > 0.01 in more than 20% samples, ii) bead number < 3 in more than 20% samples, ii) SNP probes and iv) ambiguous mapping probes^54,55^. After QC, the number of samples and probes were 153/411,585 and 155/765,201 CpG sites for adipose and muscle data, respectively. The data was then beta mixture quantile (BMIQ) normalized to adjust for type2 probe bias.

Because a considerable amount of batch effect remained from 450K and EPIC platforms in adipose tissue, we applied ComBat to minimize the effect of a platform (Supplementary Fig. 3). Methylation M-values, calculated as the log_2_ ratio between the methylated versus unmethylated probe intensities, were used in the statistical analysis^56^.

### Epigenetic age acceleration estimates

Epigenetic age for each individual was calculated from the preprocessed DNA methylation data. We used the principal component (PC) versions of the original *Hannum*^57^*, Horvath*^58^*, GrimAge*^59^ and *PhenoAge*^60^ clocks as that has been shown to remove bias caused by technical variation in certain CpGs^61^. Other epigenetic clocks applied were DunedinPACE^62^ which measures a pace of aging, as well as muscle specific epigenetic clock MEAT^63^ that was calculated for skeletal muscle tissue only. Epigenetic age acceleration measures, defined as the residuals from regressing an epigenetic age estimate on chronological age, were used in the statistical analyses.

### RNA sequencing data

Adipose tissue RNA sequencing data was available for a subset of the twins (n=80 individuals). The generation and preprocessing of the RNA-seq data has been described in detail elsewhere^11^. Briefly, RNA was extracted using AllPrep DNA/RNA/miRNA Universal Kit (Qiagen) with DNase I (Qiagen) digestion according to manufacturers’ protocol. After the calculation of RNA integrity numbers, the libraries were prepared with Illumina Stranded mRNA preparation and sequenced with Illumina HiSeq2000 platform. The data was mapped against human reference genome 38, the quality was calculated and read counts generated.

### Statistical analysis

#### Differential methylation analysis

To identify individual CpG sites associated with mtDNAq in adipose and muscle tissue, we performed an EWAS using R package *limma*^64^ that fits a linear model for each probe and computes moderated Bayes t-statistics. The models were adjusted for known biological and behavioral (age, sex, smoking status), and technical (beadchip date and row) covariates, as well as cell type proportions. In the absence of reference-based cell type deconvolution methods developed specifically for either adipose or muscle tissues, we applied EpiDISH^65^ to adjust for key known cell types: fibroblasts and epithelial cells. In addition, the adipose tissue model was adjusted for the proportion of fat cells derived from EpiDISH. The fraction of immune cells was omitted from the model due to high correlation with other cell types. The relatedness of twins in a pair was accounted for as a blocking factor in the model.

#### Associations between DNA methylation and gene expression in adipose tissue

DNA methylation is known to influence gene expression, due to which we investigated whether the identified DNA methylation sites correlated with the expression of their closest genes. We applied Pearson correlation to see to what extent DNA methylation and the expression of their respective genes were related. In addition, using linear mixed effects modeling between gene expression (outcome) and DNA methylation (predictor), and adjusting for the relatedness of the twins, we investigated how much of the variation in gene expression is explained i) by the changes in DNA methylation solely (marginal R squared) and ii) together with common familial factors shared within the twin pairs (conditional R squared).

#### Associations between obesity-related outcomes, mitochondrial DNA quantity and DNA methylation

Given that there is a well-established link between mitochondria and obesity, we were interested to explore whether the mtDNAq-associated CpG methylation was further linked to various obesity-related measures. We performed a generalized equation estimation (gee) regression using the R package *geepack*^66^ with ‘exchangeable’ correlation structure to account for within twin pair similarities. The CpG methylation was used as an outcome variable and each obesity-related outcome separately as a predictor variable. Those obesity-related variables that did not follow normal distribution were log_10_ transformed before the analysis (Supplementary Tables 3-4). The models were adjusted for age, sex and smoking, beadchip date, and row. We verified the associations between the obesity-related outcomes and mtDNAq and chose variables that exhibited statistically significant (FDR<0.05) links with both mtDNAq and CpG methylation.

#### Causal inference

To assess the potential evidence for causal relationship or common familial confounding underlying the identified associations between DNA methylation, mtDNAq and obesity-related outcomes, we applied a statistical method called ICE FALCON (Inference about Causation from Examination of FAmilial CONfounding)^15^. The method is based on regression models of observational data of related individuals, specifically twins, and assesses the changes in twin’s own and co-twin’s regression coefficient from with and without adjusting the counterparts’ predictor variables (Models 1-3 below). We restricted the ICE FALCON analysis for the complete MZ pairs in our cohort.

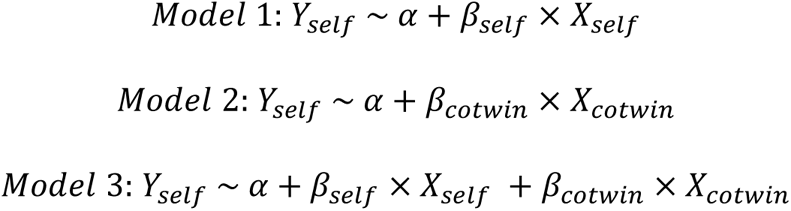

Briefly, if the observed cross-twin cross-trait association (β_cotwin_) diminishes after adjusting for within-individual association it suggests a causal effect from the predictor variable to the outcome. Conversely, if the β_cotwin_ appears only when conditioning on the within-individual association, it indicates a causal relationship from the outcome to the predictor. If both the β_cotwin_ and the β_self_ decrease after conditioning on each other, it indicates familial confounding (genetic or environmental) between the predictor and outcome. We examined β_cotwin_ with and without conditioning on within-individual association using generalized equation estimation in R package *geepack*^62^ using ‘exchangeable’ correlation structure for the twin pairs. All analyses were adjusted for age, sex and smoking. Technical covariates of beadchip date and row were added as covariates to adjust analyses including DNA methylation data.

Additionally, the ICE FALCON between mtDNAq and DNA methylation sites were further adjusted for BMI. We then calculated the changes in regression coefficients, for which we estimated standard errors of changes with non-parametric bootstrapping generating 1,000 datasets having the original sample size. The models were then reversed, e.g. using the previous X variable as Y variable to gain additional evidence on the causal pathway.

## Acknowledgements

The authors thank all the study participants. We also acknowledge the computational resources of the Institute for Molecular Medicine Finland (FIMM) Technology Center. S.Lu. was involved in this research while affiliated with the Institute for Molecular Medicine Finland (FIMM) and is currently affiliated with Nightingale Health Plc.

## Author contributions

A.He. and M.O. designed the study. A.He. performed the data analyses, visualized the data and wrote the first draft of the manuscript. K.H.P., S.H., M.O. and J.Ka. collected and generated the data. A.He. and S.H.T.L. preprocessed the omics data. J.Ku., A.Ha. and J.L. performed the MRI and P-H.G. the DEXA imaging. P.P. generated the RNA sequencing data. V.F.C.E. and S.Li. developed and assisted on the ICE FALCON analysis. M.O. and S.Lu. supervised the work. All authors critically commented and edited the final version of the manuscript.

## Ethics

The Ethics Committee of the Hospital District of Helsinki and Uusimaa approved the protocols of this FTC substudy data collections, and all participants provided their written informed consent. The authors assert that all procedures contributing to this work comply with the ethical standards of the relevant national and institutional committees on human experimentation and with the Declaration of Helsinki.

## Data availability

RNA sequencing data is part of the ‘Twin Study’ and deposited with the Biobank of the Finnish Institute for Health and Welfare (https://thl.fi/en/research-and-development/thl-biobank/for-researchers/sample-collections/twin-study). For details on accessing the data, see: https://thl.fi/en/research-and-development/thl-biobank/for-researchers/application-process. The rest of the FTC data is not publicly available due to the restrictions of informed consent. However, the FTC data is available through the Institute for Molecular Medicine Finland (FIMM) Data Access Committee (DAC) for authorized researchers who have IRB/ethics approval and an institutionally approved study plan.

