## Supplementary Tables and Figures for "Twin pair analysis uncovers novel links between DNA methylation, mitochondrial DNA quantity and obesity"

**Supplementary Materials**

Supplementary Table 1. Epigenetic age estimates of the study cohort

| Epigenetic clock | Tissue | Mean (SD) , years* |
|---|---|---|
| **Horvath clock** | Adipose | -0.2 (-8.7–7.7) |
| | Muscle | -0.3 (-7.6–6.2) |
| **Hannum clock** | Adipose | -0.2 (-8.0–6.5) |
| | Muscle | -0.1 (-5.3–5.0) |
| **GrimAge** | Adipose | -0.2 (-5.8–4.1) |
| | Muscle | -0.1 (-3.7–4.0) |
| **PhenoAge** | Adipose | -0.2 (-11.5–11.8) |
| | Muscle | -0.2 (-8.1–4.6) |
| **DunedinPACE*** | Adipose | 1.7 (1.5–2.0) |
| | Muscle | 1.4 (1.2–1.5) |
| **MEAT** | Muscle | 0.2 (-8.5–7.3) |

*** unit: years/calendar year**

Supplementary Table 2. Characteristics of the monozygotic twin pairs used in ICE FALCON analysis of SH3BP4 methylation, mtDNA quantity and obesity-related traits in adipose tissue.

| Variable | Heavier co-twin (mean) | Leaner co-twin (mean) | Difference (Heavier-Leaner) | P-value** | N (MZ twin pairs) |
|---|---|---|---|---|---|
| cg19998400* (M-value) | 0.82 | 0.79 | 0.03 | 8.0E−05 | 68 |
| mtDNAq* | −0.06 | 0.02 | −0.08 | 7.7E−05 | 71 |
| Weight (kg) | 91.60 | 77.84 | 13.76 | 5.4E−18 | 71 |
| BMI (kg/m2) | 31.93 | 27.32 | 4.61 | 4.6E−19 | 71 |
| Waist circumference (cm) | 104.38 | 91.96 | 12.42 | 3.6E−17 | 64 |
| WHR | 0.93 | 0.90 | 0.04 | 8.9E−05 | 33 |
| Fat (%) | 40.65 | 35.33 | 5.32 | 1.4E−13 | 71 |
| Fat (kg) | 38.36 | 28.79 | 9.57 | 3.0E−14 | 71 |
| Fat-free mass (kg) | 51.14 | 47.60 | 3.55 | 1.4E−07 | 71 |
| Intra-abdominal fat (cm3) | 1300.57 | 625.16 | 675.42 | 7.7E−04 | 21 |
| Subcutaneous fat (cm3) | 5933.65 | 3739.47 | 2194.18 | 3.8E−05 | 21 |
| Adipocyte volume (mm3) | 615 | 457 | 159 | 7.3E−08 | 60 |
| Liver fat (%) | 4.15 | 1.60 | 2.55 | 9.9E−04 | 42 |
| Total cholesterol (mmol/l) | 4.82 | 4.73 | 0.09 | 0.47 | 71 |
| HDL (mmol/l) | 1.47 | 1.64 | −0.18 | 1.7E−04 | 71 |
| LDL (mmol/l) | 3.12 | 2.91 | 0.22 | 0.07 | 71 |
| Triglycerides (mmol/l) | 1.12 | 0.97 | 0.16 | 0.01 | 71 |
| hsCRP (mg/l) | 3.33 | 2.31 | 01.02 | 0.02 | 64 |
| Adipsin (ug/l) | 1.21 | 1.14 | 0.07 | 0.12 | 21 |
| Adiponectin (ng/l) | 2903.00 | 3694.00 | -791.00 | 7.7E−04 | 21 |
| ALAT (U/l) | 29.20 | 24.93 | 4.27 | 0.03 | 71 |
| ASAT (U/l) | 28.94 | 27.27 | 1.68 | 0.40 | 71 |
| BP (systolic) (mmHg) | 134 | 134 | 0 | 0.74 | 69 |
| BP (diastolic) (mmHg) | 81 | 79 | 2 | 0.03 | 69 |
| Fasting glucose (mmol/l) | 5.82 | 5.55 | 0.27 | 0.11 | 69 |
| Fasting insulin (mU/l) | 9.50 | 7.28 | 2.22 | 3.0E−03 | 67 |
| HOMA-IR | 2.52 | 1.81 | 0.71 | 2.5E−03 | 64 |
| Matsuda index | 5.58 | 7.76 | −2.18 | 7.8E−05 | 59 |
| Leisure-time physical activity | 2.91 | 2.92 | −0.02 | 0.86 | 65 |
| Sports activity | 2.59 | 2.77 | −0.18 | 0.12 | 66 |

| | | | | | |
|---|---|---|---|---|---|
| Work activity | 2.45 | 2.44 | 0.02 | 0.91 | 67 |
| Total activity | 7.98 | 8.17 | −0.19 | 0.29 | 63 |
| EAAHorvath* | 0.14 | −0.47 | 0.61 | 0.02 | 68 |
| EAAHannum* | 0.35 | −0.49 | 0.84 | 5.5E−04 | 68 |
| EAAGrim* | −0.43 | −0.03 | −0.39 | 0.05 | 68 |
| EAAPheno* | 0.26 | −0.34 | 0.60 | 0.08 | 68 |
| DunedinPACE* | 1.70 | 1.69 | 0.00 | 0.80 | 68 |

* Estimates derived from adipose tissue

** Paired t-test (two-tailed)

ALAT = Alanine aminotransferase; ASAT = Aspartate aminotransferase; BMI = body mass index; DZ = dizygotic; EAA = Epigenetic Age Acceleration; HOMA-IR = Homeostatic model assessment for insulin resistance; hsCRP = high-sensitivity C-reactive protein; mtDNAq = mitochondrial DNA quantity; MZ = monozygotic; WHR = waist-to-hip ratio

Supplementary Table 3. Results from the causal inference using ICE FALCON between the expression and methylation of SH3BP4 and DHRS3 (n=40 monozygotic twin pairs).

| Formula | Coefficient | Model 1 | | | Model 2 | | | Model 3 | | | Change | |
|---|---|---|---|---|---|---|---|---|---|---|---|---|
| | | Est | SE | p.val | Est | SE | p.val | Est | SE | p.val | Est | p.val |
| SH3BP4 ~ cg19998400 | Bself | 0.261 | 0.047 | 2.0E-08 | | | | 0.267 | 0.047 | 1.3E-08 | 0.006 | 0.871 |
| | Bcotwin | | | | -0.112 | 0.064 | 0.078 | 0.014 | 0.066 | 0.833 | 0.126 | 0.134 |
| cg19998400 ~ SH3BP4 | Bself | 0.652 | 0.186 | 4.7E-04 | | | | 0.717 | 0.163 | 1.0E-05 | 0.065 | 0.494 |
| | Bcotwin | | | | 0.031 | 0.162 | 0.846 | -0.225 | 0.173 | 0.193 | -0.256 | 0.299 |
| DHRS3 ~ cg17468563 | Bself | -0.643 | 0.156 | 3.7E-05 | | | | -0.657 | 0.146 | 6.7E-06 | -0.014 | 0.822 |
| | Bcotwin | | | | -0.050 | 0.191 | 0.793 | 0.075 | 0.165 | 0.647 | 0.125 | 0.532 |
| cg17468563 ~ DHRS3 | Bself | -0.296 | 0.072 | 3.7E-05 | | | | -0.306 | 0.071 | 1.5E-05 | -0.010 | 0.696 |
| | Bcotwin | | | | -0.008 | 0.078 | 0.916 | -0.059 | 0.055 | 0.280 | -0.051 | 0.581 |

Supplementary Table 4. Associations between adipose tissue SH3BP4 methylation and obesity-related traits.

| Outcome | Coef | SE | P-value | FDR | N |
|---|---|---|---|---|---|
| Weight | 0.246 | 0.070 | 4.1E-04 | **0.002** | 136 |
| BMI | 0.283 | 0.078 | 2.9E-04 | **0.001** | 136 |
| Waist | 0.300 | 0.065 | 4.0E-06 | **3.5E-05** | 122 |
| WHR | 0.123 | 0.062 | 0.050 | 0.079 | 64 |
| Fat % | 0.290 | 0.062 | 2.7E-06 | **4.7E-05** | 136 |
| Fat (kg) | 0.318 | 0.077 | 3.4E-05 | **2.3E-04** | 136 |
| Fat-free mass | 0.051 | 0.039 | 0.190 | 0.247 | 136 |
| Ia. Fat[a] | 0.458 | 0.099 | 3.6E-06 | **4.1E-05** | 42 |
| Subcut. Fat[a] | 0.352 | 0.086 | 4.5E-05 | **2.6E-04** | 42 |
| Adipocyte volume | 0.202 | 0.079 | 0.010 | **0.022** | 114 |
| Liver fat %[a] | 0.391 | 0.080 | 1.1E-06 | **3.8E-05** | 82 |
| Total cholesterol | 0.027 | 0.088 | 0.761 | 0.807 | 136 |
| HDL | -0.189 | 0.068 | 0.006 | **0.015** | 136 |
| LDL | 0.059 | 0.092 | 0.523 | 0.631 | 136 |
| Triglycerides[a] | 0.222 | 0.090 | 0.013 | **0.028** | 136 |
| hsCRP[a] | 0.320 | 0.081 | 7.2E-05 | **3.6E-04** | 122 |
| Adipsin | 0.058 | 0.146 | 0.694 | 0.759 | 42 |
| Adiponectin | -0.275 | 0.089 | 0.002 | **0.006** | 42 |
| ASAT[a] | 0.033 | 0.077 | 0.672 | 0.784 | 136 |
| ALAT[a] | 0.095 | 0.088 | 0.282 | 0.353 | 136 |
| BP (systolic) | 0.089 | 0.057 | 0.122 | 0.165 | 132 |
| BP (diastolic) | 0.206 | 0.063 | 0.001 | **0.003** | 132 |
| Fasting glucose[a] | 0.165 | 0.074 | 0.025 | **0.046** | 132 |
| Fasting insulin[a] | 0.233 | 0.097 | 0.016 | **0.032** | 128 |
| HOMA-IR[a] | 0.250 | 0.095 | 0.009 | **0.021** | 122 |
| Matsuda[a] | -0.303 | 0.089 | 0.001 | **0.002** | 112 |
| LTPA | -0.246 | 0.092 | 0.008 | **0.019** | 124 |
| Sports activity | -0.178 | 0.088 | 0.042 | 0.074 | 126 |
| Work activity | 0.026 | 0.097 | 0.793 | 0.816 | 128 |
| Total activity | -0.169 | 0.086 | 0.049 | 0.082 | 120 |
| EAAHorvath | 0.014 | 0.070 | 0.837 | 0.837 | 136 |
| EAAHannum | 0.134 | 0.070 | 0.055 | 0.080 | 136 |
| EAAGrim | -0.116 | 0.069 | 0.090 | 0.126 | 136 |
| EAAPheno | 0.026 | 0.063 | 0.678 | 0.766 | 136 |
| DunedinPACE | -0.156 | 0.080 | 0.053 | 0.081 | 136 |

[a] Log10 transformed

Supplementary Table 5. Associations between adipose tissue mtDNA quantity and obesity-related traits.

| Outcome | Coef | SE | P-value | FDR | N |
|---|---|---|---|---|---|
| Weight | -0.346 | 0.096 | 3.2E-04 | **0.001** | 142 |
| BMI | -0.340 | 0.095 | 3.5E-04 | **0.001** | 142 |
| Waist | -0.320 | 0.095 | 0.001 | **0.002** | 128 |
| WHR | -0.175 | 0.095 | 0.066 | 0.096 | 66 |
| Fat % | -0.239 | 0.073 | 0.001 | **0.002** | 142 |
| Fat (kg) | -0.357 | 0.091 | 9.4E-05 | **0.001** | 142 |
| Fat-free mass | -0.184 | 0.061 | 0.002 | **0.006** | 142 |
| Ia. Fat[a] | -0.578 | 0.117 | 7.5E-07 | **2.6E-05** | 42 |
| Subcut. Fat[a] | -0.464 | 0.105 | 1.1E-05 | **1.2E-04** | 42 |
| Adipocyte volume | -0.322 | 0.085 | 1.6E-04 | **0.001** | 120 |
| Liver fat %[a] | -0.366 | 0.123 | 0.003 | **0.006** | 84 |
| Total cholesterol | 0.076 | 0.087 | 0.382 | 0.432 | 142 |
| HDL | 0.324 | 0.089 | 2.7E-04 | **0.001** | 142 |
| LDL | -0.062 | 0.092 | 0.505 | 0.535 | 142 |
| Triglycerides[a] | -0.159 | 0.108 | 0.142 | 0.184 | 142 |
| hsCRP[a] | -0.194 | 0.109 | 0.074 | 0.103 | 128 |
| Adipsin | -0.229 | 0.100 | 0.023 | **0.040** | 42 |
| Adiponectin | 0.357 | 0.098 | 2.7E-04 | **0.001** | 42 |
| ALAT[a] | -0.095 | 0.099 | 0.337 | 0.393 | 142 |
| ASAT[a] | -0.091 | 0.092 | 0.322 | 0.389 | 142 |
| BP (systolic) | 0.025 | 0.074 | 0.739 | 0.761 | 138 |
| BP (diastolic) | -0.163 | 0.085 | 0.057 | 0.086 | 138 |
| Fasting glucose[a] | -0.097 | 0.085 | 0.258 | 0.322 | 138 |
| Fasting insulin[a] | -0.296 | 0.103 | 0.004 | **0.009** | 134 |
| HOMA-IR[a] | -0.355 | 0.093 | 1.4E-04 | **0.001** | 128 |
| Matsuda[a] | 0.345 | 0.095 | 2.8E-04 | **0.001** | 118 |
| LTPA | 0.183 | 0.094 | 0.051 | 0.080 | 130 |
| Sports activity | 0.260 | 0.101 | 0.010 | **0.020** | 132 |
| Work activity | -0.068 | 0.092 | 0.458 | 0.501 | 134 |
| Total activity | 0.194 | 0.093 | 0.038 | 0.063 | 126 |
| EAAHorvath | -0.176 | 0.073 | 0.016 | **0.030** | 136 |
| EAAHannum | -0.273 | 0.072 | 1.4E-04 | **0.001** | 136 |
| EAAGrim | 0.145 | 0.091 | 0.110 | 0.148 | 136 |
| EAAPheno | -0.252 | 0.054 | 3.2E-06 | **5.5E-05** | 136 |
| DunedinPACE | -0.004 | 0.094 | 0.964 | 0.964 | 136 |

[a] Log10 transformed

Supplementary Figure 1. Scatter plot representing the regression coefficients from EWAS on mtDNA quantity in adipose tissue (X) and skeletal muscle (Y).

Supplementary Figure 2. The amount of methylation probe quantile variation unexplained by the control probe principal components under 10-fold cross validation in (a) adipose tissue and (b) skeletal muscle. The number of principal components which the probe quantiles were normalized with is indicated in the plots.
M = methylated probes; U = unmethylated probes

Supplementary Figure 3. First four principal components of the adipose tissue DNA methylation data indicated by beadchip type (450K/EPIC), (a) before and (b) after adjustment with *ComBat*.

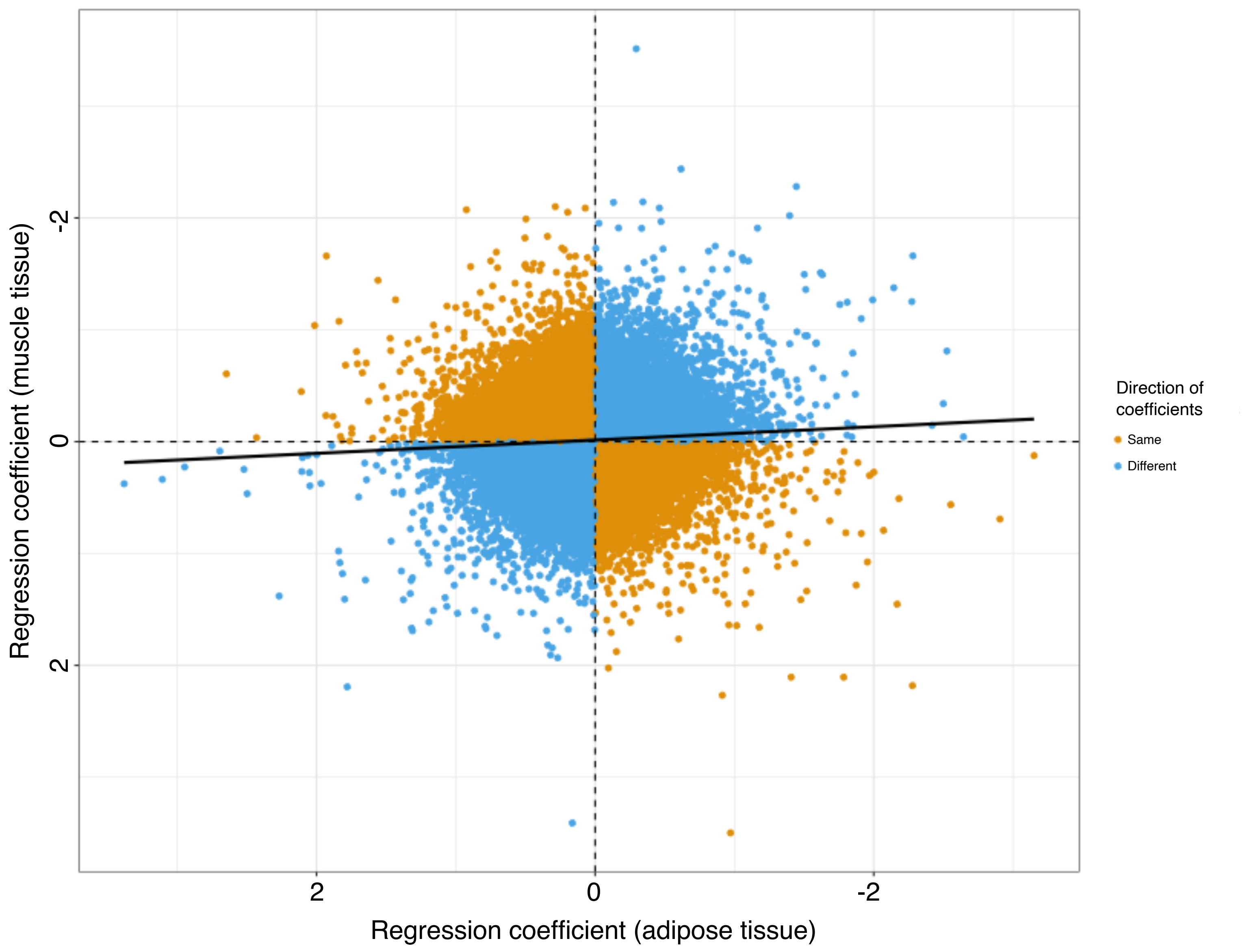

Regression coefficient (muscle tissue)
-2
0
2
Regression coefficient (adipose tissue)
2
0
-2
Direction of coefficients
Same
Different

a

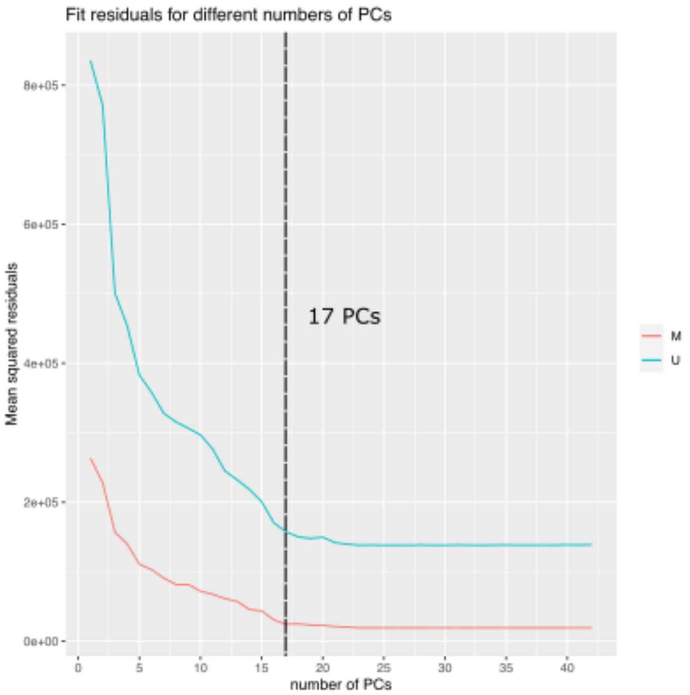

Fit residuals for different numbers of PCs
Mean squared residuals
8e+05
6e+05
4e+05
2e+05
0e+00
0
5
10
15
20
25
30
35
40
number of PCs
17 PCs
M
U


b

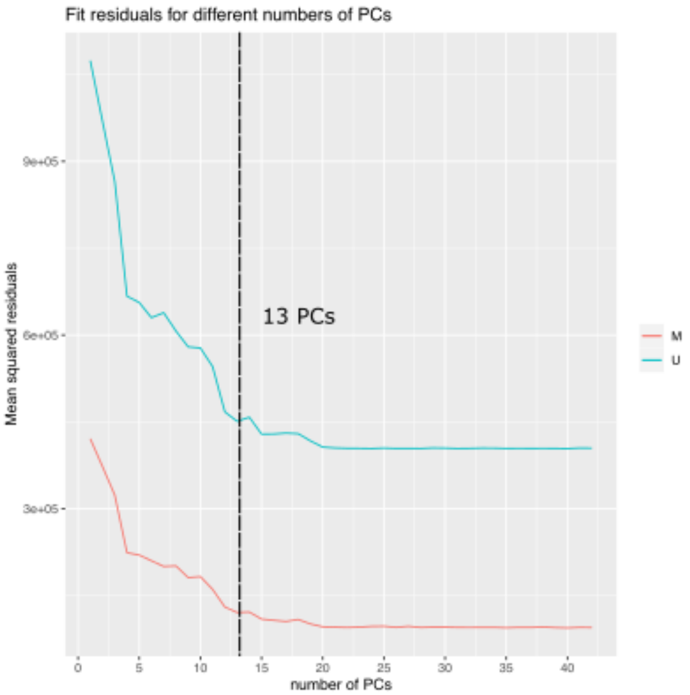

Fit residuals for different numbers of PCs
Mean squared residuals
9e+05
6e+05
3e+05
0
5
10
15
20
25
30
35
40
number of PCs
13 PCs
M
U

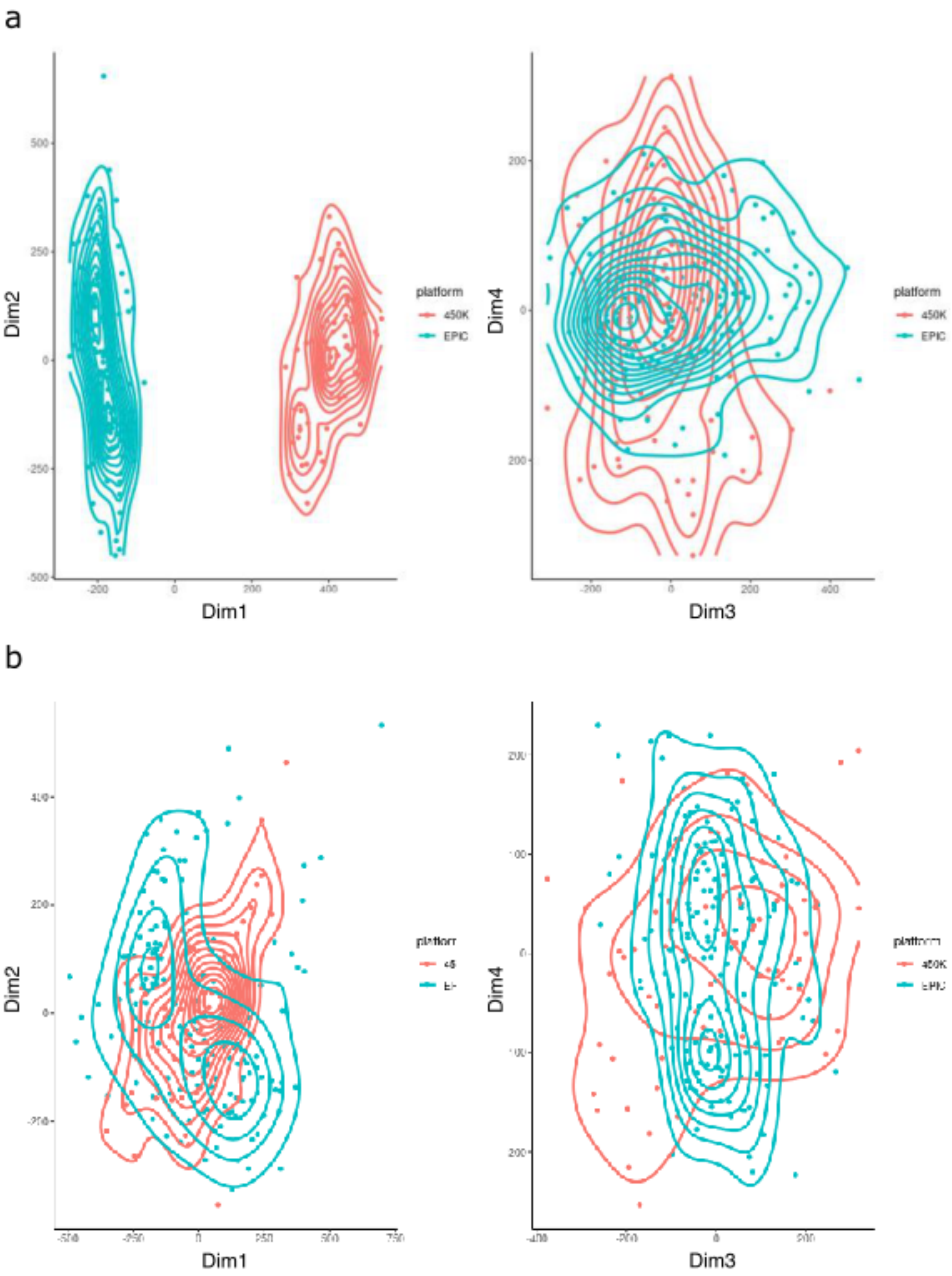
a
b
Dim1
Dim2
Dim3
Dim4
platform
450K
EPIC
